# Semantic Encoding in Medical LLMs for Vocabulary Standardisation

**DOI:** 10.1101/2025.06.16.25329716

**Authors:** Samuel Mainwood, Aashish Bhandari, Sonika Tyagi

## Abstract

High-quality, standardised medical data availability remains a bottleneck for digital health and AI model development. A major hurdle is translating noisy free text into controlled clinical vocabularies, aiming for harmonisation and interoperability, especially when source datasets are inconsistent or incomplete. We benchmark domain-specific encoder models against general LLMs for semantic-embedding retrieval using minimal vocabulary building blocks and test several prompt techniques. We also try prompt augmentation with LLM-generated differential definitions. We tested these prompts on open-source Llama and medically fine-tuned Llama models to steer their alignment toward accurate concept assignment across multiple prompt formats. Domain-tuned models consistently outperform general models of the same size in retrieval and generative tasks. However, performance is sensitive to prompt design and model size, and the benefits of adding LLM-generated context are inconsistent. While newer, larger foundation models are closing the gap, today’s lightweight open-source generative LLMs lack the stability and embedded clinical knowledge needed for deterministic vocabulary standardisation.

## 1 Introduction

For a system to be successfully integrated and approved for use within healthcare, it must demonstrate rigid efficacy, reliability and interpretability at a higher burden than other domains [16]. One of the significant barriers to developing these clinical models is the quantity, quality, and availability of data [39, 48]. When developing general domain large language models (LLM), large companies can send public releases to be broken and iron out any unwanted quirks. Healthcare does not have that same luxury, as these faults can result in adverse patient outcomes, so the burden of consistency and performance at roll-out is much higher. Electronic Health Records (EHR) is a term that refers to clinical databases with a wealth of patient information. This information includes both structured and unstructured data. Structured data typically refers to organised tabular data in the form of numerical and categorical information. Unstructured data, as the name implies, can take any form and often consists of documents of text, images and many other forms that have not historically been easy to use in computational problems. Improving data quality and availability is one facet. Another challenge is harmonising diverse sources into usable and consistent datasets for machine learning applications [38]. “Medical concept mapping” refers to converting medical information into a controlled vocabulary. This study defines medical concepts as entities that represent medical terminology related to disease, diagnoses, symptoms, treatments, interventions, and all other information that could characterise the semantic meaning of a medical concept. This process aims to facilitate interoperability through consistency in data whilst maintaining the semantic essence of a term, agnostic of context. One of the ways that medical concept data is stored is in the form of an ontology [19]. An ontology is a knowledge graph that holds semantic information in the form of classes, including unique identifying features [19, 41]. This study will explore ontology mapping from the OAEI BioML track as a proxy for standardising clinical terminology. The automation of a reliable medical ontology conversion system is an essential step in addressing medical data standardisation.

## 2 Background

The BioML track of the Ontology Alignment Evaluation Initiative (OAEI) is notoriously challenging; methods effective in other ontology matching tasks often fail to generalise well here due to the distinct complexities inherent in biomedical ontologies [2, 32]. For instance, while large-scale biomedical ontologies such as SNOMED CT have richly structured semantic relationships, simpler classification systems like ICD-10 [45], ICD-11 [46], and the UK Biobank dataset lack such extensive semantic structures [37]. These simpler ontologies typically present as tabular datasets without hierarchical relationships or entity types, limiting the applicability of more sophisticated ontology matching approaches [37].

Several tools and frameworks have addressed medical vocabulary standardisation. USAGI, an open-source tool, relies primarily on information retrieval and has successfully mapped UK Biobank medical data to SNOMED CT [34, 37, 38]. Although accurate within their validated domain, rule-based methods such as Ontoserver, lack dataset flexibility and generalisability [28, 40]. MedCAT, another open-source toolkit, combines traditional natural language processing (NLP) techniques with manual expert involvement but relies on iterative reinforcement learning to maintain performance in evolving clinical contexts [20]. EHR-QC further extends traditional NLP through ensemble methods, integrating tools like MedCAT, fuzzy matching and reverse indexing, achieving good results validated against simpler datasets such as the UK Biobank [38]. Despite traditional NLP methods initially setting benchmarks, recent advancements using transformer-based LLMs have significantly surpassed earlier techniques in medical NLP tasks [49]. Medical and domain-specific models tend to outperform their generalist counterparts on medical tasks regarding size and training volume size relative to performance [1, 22, 24, 25, 49]. Nonetheless, LLM-based approaches in ontology matching have faced difficulties despite competitive performance in generalist tracks [3, 31, 32]. Conversely, the retrieveidentify-prompt pipeline proposed by Taboada *et al*. demonstrated that high-quality embeddings alone could yield competitive results without complex decoder LLM frameworks, despite not exploring domain-specific embeddings or nuanced prompting methods [42]. Additionally, some models have achieved high benchmarks through ensemble models that include embedding, graph matching and ontology repair, but these methods are incompatible with simpler datasets [12, 13, 17, 28, 36]. The most comprehensive prompt evaluation study did not evaluate on the BioML track [15]. Common evaluation challenges include sparse ontology intersections and inconsistencies in curated “gold-standard” datasets, compounded by non-named entities lacking clinical relevance and complicating NLP-based methods [14]. Given these complexities, and considering real-world biomedical datasets tend to be simpler and less granular, focusing purely on label-based ontology matching emerges as a practically justified approach for this study. This more straight-forward label-based strategy aligns with realistic biomedical use cases.

## 3 Methodology

Tokenisation is the first step in processing clinical documents that converts a document into character or word representations called “tokens”. Tokens can then be processed into numerical representations through encoding and storing them as embedding vectors [43]. Tokenisation combined with the transformer architecture [10] enables a reinforcement-style training approach by feeding the model millions of tokens and having the model slowly settle on embeddings or vectors that would align with token prediction. This resulted in a compact, efficient model with the most robust semantic embeddings. The resulting direction from a tokenised and vectorised document holds calculable meaning. The resulting vectors can then be used to calculate their similarity, resulting in scalable semantic similarity search.

### 3.1 Concept Matching Overview

The task involves identifying one-to-one mappings between items in a source vocabulary and concepts in a controlled vocabulary. For terminology clarity, this paper defines a confirmed match as cases where a model identifies two concepts from distinct vocabularies representing the same underlying medical concept. A non-match is a case in which the model identifies no appropriate concept within the controlled vocabulary corresponding to the source concept.

### 3.2 Semantic Embedding Matching Algorithm

To match embeddings, this paper used Euclidean distance and Cosine Similarity as a comparison. High-quality embeddings should achieve similar results for retrieval across all three matching algorithms if the embeddings have good alignment. Cosine similarity was preferred for its ability to match with a relative metric instead of the absolute metric provided by the Euclidean distance. To define non-matches, Cosine similarity enables thresholding strategies that are not possible using Euclidean distance. This is explored across five OAEI ontologies. The strength of simple embedding matchings is their robustness, reliability, and simplicity, which are much more explainable and controlled than generative LLM models. The challenge with standalone use is that there is always a confirmed match retrieved. The ability to assign non-matches whilst leveraging the high-performance of retrieval methods is paramount in defining the best system. The benchmark algorithm was to consider a simple threshold to assign non-matches based on the decay of F1 across lowered thresholds. Other studies have used a 0.9 threshold as a filter for Decoder LLM queries [2, 3, 42]. The quadratic complexity *O* (*n*^2^) of ontology matching, compounded by the computational demands of LLMs and the infeasibility of storing or re-calculating embeddings in memory, made early list-based approaches (e.g., requiring ∼ 4 hours per match) untenable. By adopting scalable vector databases like ChromaDB (disk-based) [8] and FAISS (RAM-based) [21], integrated via LangChain [21], the search complexity was reduced to *O* (2*n*) through constant-time nearest-neighbour lookups. Customising feature embeddings for vocabulary standardisation does not increase model complexity but can impact embedding quality. While adding context to labels may seem beneficial, prior work [3] and early experiments found it degrades performance by overwhelming the embedding space. We used a variety of domain-specific models, including ClinicalT5’s encoder [24] and a series of domain-specific BERT models with varying training sizes [1, 29, 48]. We also compared self-aligned domain-specific models that have undergone targeted vocabulary pre-training [22, 25].

### 3.3 Identifying Non-Matches through LLM

Retrieval augmented generation (RAG) refers to a secondary component in an LLM model that is highly adept at returning relevant documents and information for the core model before using a decoder. The major component of this study explored the use of the domain-specific semantic embeddings as a RAG tool to generate higher-quality, precise prompts for generative LLMs. Instead of retrieving the highest match, the highest K-matches are retrieved, and then the results can be given to an LLM to avoid using an arbitrary threshold. RAG allows the user to maintain a minimal problem size and significantly constrains the output space to prevent LLM hallucination.

### 3.4 Prompting Techniques

Prompt content is among the most significant challenges in generative LLM processes. Prompting aims to align the model’s output with the intended task. Prompting is an infinite space, and whilst there is some theoretical basis for what a “good” prompt requires [23, 30], knowledge of it is still growing. High-level prompt templates exist, but no prompt template in published studies has demonstrated conclusively better performance. The prompts used in this study, when using LLM models, are available in the appendix. Whilst encoding models have a deterministic output that absorbs natural language as text to compute a vector, generative models have increasingly complex hyperparameters that can make it challenging to control their output.

#### 3.4.1 Chattiness and Common LLM Weaknesses

Chattiness refers to generating verbose or overly detailed outputs in response to prompts. This behaviour poses a reliability issue in automated pipelines, especially those requiring structured or binary outputs. Decoder-only models, particularly with fewer parameters, often fail to constrain their output despite explicit, minimal instructions. Excess tokens or unexpected formatting reduce usability even when the generated content is semantically correct. Therefore, this pipeline penalises any deviation from the expected output format, such as added explanations or uncertainty phrases, by marking the response as a non-match. Prompt refinement was performed iteratively on a small sample dataset until the model reliably adhered to the task description and expected output format. Common issues observed included the model selecting multiple target codes for a single input concept, particularly when the model interpreted an input as equally similar to multiple target codes. Additionally, quantised LLaMA models tended to erroneously return the input code as the chosen match, necessitating further prompt adjustments. Consequently, due to these instabilities, the final retrieval step treated any output that deviated from the specified format or presented invalid candidate codes as a non-match, thereby preserving output consistency and integrity. In early experiments with Llama3.1, a model was asked to give a “confidence” score, but this did not appear to be a deterministic number and instead made the model “chattier” by vocalising the need to explain the score number despite explicit instructions otherwise.

#### 3.4.2 LLM Hyperparameters

Quantisation is a model compression technique that reduces the memory footprint and computational demands of LLMs by representing weights and activations using low-bit integers instead of full floating precision numbers, whilst preserving performance [18]. In this study, all quantised models were obtained via Ollama [35], which distributes pre-quantised models optimised for local inference. The relevant models will indicate quantisation intensity. This study used a temperature of 0.0, or another hyperparameter alternative that uses a deterministic decoder output. With something as rigid as standardisation, a higher temperature may allow freedom for niche codes to be mapped in “creative” ways. However, this compromises the already questionable reliability and consistency of generative LLMs, and by setting the temperature to 0, the model should return the same consistent and reliable outputs in response to the same inputs.

#### 3.4.3 Multi-Choice vs Binary Questioning

Most decoder models are released with evaluation on question-answering tasks (QA) [5– 7, 11, 26, 47, 49]. As such, it is logical that the next step is reframing label generation as a question. It is infeasible to list all codes from large ontologies within a prompt context window, but using a RAG tool enables a much smaller, targeted set of *top* − *k* to be retrieved and presented. Multiple-choice questioning shows the input term with its *top* −*k* candidate codes and asks the model to return the best match, or if there are no appropriate matches, returns None. Early smoke tests with Q4_K_M Llama 3.1 showed that prompts had to be iteratively tightened to curb “chatty” responses and also prevent the model from echoing the input code. This technique allows more candidates to be provided to an LLM at lower computational cost through a single prompt, but increases prompt complexity, making it harder to adjust effectively. Binary questioning for ontology matching gives the model an input prompt with a potential match, asking true/false or another binary outcome. The simplicity of this prompt makes it easier to align the decoder output. There are some emerging challenges with this approach. Firstly, asking only for the best match excludes the demonstrated improvement for the retrieval models at *k* = 5 and *k* = 10 matches without adding further algorithmic elements.

#### 3.4.4 Context Enhancement using LLM Generated Differential Definitions Separation

Generative LLMs improve their task alignment with additional context. Due to the lack of ontology homogeneity, the task is to build prompt context without additional information. One persistent limitation of current artificial intelligence models is their inability to generate or consider counterfactual scenarios or abstain from responding. This manifests as a failure to recognise alternative, potentially superior matches within the same ontology beyond the immediate best apparent match. For instance, consider the Foundational Model of Anatomy (FMA) [4] code “44620” (Medial Meniscus), which may appear as the closest match to the SNOMED [44] input “74135004” (Meniscus structure of joint). While contextually plausible, other closer semantic matches within the ontology remain unconsidered by models that lack context. Without additional context, models cannot infer or recognise superior matching possibilities that a human might naturally consider. The proposed method first prompts the LLM to consider an input concept alongside its *top* −*K* semantically similar candidates, which are derived using the embedding matching retrieval method from within the same ontology. The decoder model is instructed to generate a definition that defines but also explicitly differentiates the input from these candidates. The underlying hypothesis is that instructing the LLM in this targeted manner enables it to produce distinct keywords or phrases that effectively adjust the semantic embedding vectors within the LLM model. Consequently, this definitional differentiation provides critical contextual cues that enhance the model’s ability to distinguish between similar medical concepts and infer the existence of similar concepts within the same ontology, thus improving match selection precision. By leveraging the generative capabilities of the LLM, the method removes subjective human biases that might influence manual definitions, aiming instead to generate precise, objective, and consistent definitions that may improve context in the subsequent LLM. A robust generative LLM approach can also avoid data bias in threshold matching. The resulting differential definitions produced by the LLM are then systematically stored for later access in the pipeline to be accessed when building prompts for negative filtering. The rationale is that using the same model to generate differential definitions produces language that the same generative model should understand downstream. A sample prompt for differential definition building, multi-choice questioning and true-false questioning can be found in the appendix.

#### 3.4.5 Datasets

The Ontology Datasets used in this study are from the OAEI 2024 Machine-Learning Friendly Datasets [14]. These included pruned and cleaned ontologies with 8 Ontologies (one ontology has three sub-ontologies), divided into five matching tasks [33]

#### 3.4.6 LLM Sampling Datasets

This study used a smaller sample dataset to balance computational availability while maintaining a reliable and holistic model performance evaluation. 200 random positive and 200 random negative items were taken from each input dataset to create five datasets of 400 items across the 5 OAEI tracks.

## 4 Results

### 4.1 Self-Aligned Domain-Specific Models Outperform in Retrieval

Table 1 demonstrates that self-aligned embedding retrievers topped every RAG benchmark at k = 5 and k = 10, and both self-aligned models had an impressive k=1. SAPPMBert is not OAEI-competitive, yet a blanket 0.90 similarity cut-off still gave mean F1 = 0.674 and precision = 0.762. Figure 1 highlights this decay across a lowered matching threshold. Such fixed thresholds remain fragile: score distributions differ sharply across ontology pairs. For instance, as demonstrated in Table 3, most SNOMED-Pharm to NCIT-Pharm alignments sit above 0.90 similarity, whereas many MONDO-OMIM to ORDO links do not. Even so, a strong, domain-tuned retriever can shoulder much of the alignment burden, and domain-specific pretraining usually outperforms general-domain retrieval methods. Table 2 demonstrates a very marginal increase in performance using Euclidean distance, but as highlighted previously, the marginal performance difference does not justify this inflexible method.

**Table 1:** Average Cosine-Distance Retrieval Performance Across Datasets. Performance is reported using Hits@K metrics (*K* = 1, 5, 10). It indicates the proportion of times the correct item appears in the top-*K* retrieved results.

| Model | Hits@1 | Hits@5 | Hits@10 |
| --- | --- | --- | --- |
| ClinicalBERT [1] | 0.578 | 0.690 | 0.718 |
| ClinicalT5 [24] | 0.553 | 0.643 | 0.670 |
| GatorTronBERT [48] | 0.524 | 0.615 | 0.646 |
| SAPBIOBERT [22] | 0.842 | **0.910** | **0.926** |
| SAPPMBERT [25] | 0.852 | **0.921** | **0.938** |
| UMLSBERT [29] | 0.602 | 0.713 | 0.745 |
| Ada OpenAI Retrieval Average [3] | – | 0.8462 | 0.8792 |

**Table 2:** Comparison of Cosine Similarity vs Euclidean Distance in Label Matching. Performance is measured using Hits@K metrics (*K* = 1, 5, 10) and Mean Reciprocal Rank (MRR). MRR gives a balanced measure of Hits@K, favouring returning the desired document at a higher rank [9]. These metrics show the effectiveness of each distance metric in retrieving the correct label. The higher value indicates better matching performance.

| Metric | Dataset | Hit @ 1 | Hit @ 5 | Hit @ 10 | MRR |
| --- | --- | --- | --- | --- | --- |
| Cosine | Average | 0.7778 | 0.8934 | 0.9100 | 0.8544 |
| Euclidean | Average | 0.7812 | 0.8986 | 0.9158 | 0.8586 |

**Table 3:** Baseline Raw with 0.90 Threshold Cut-off. Precision, Recall, and F1 scores are reported for each dataset using a fixed confidence threshold of 0.90. These metrics evaluate label matching quality. The bottom row shows the macro-average across all datasets.

| Dataset | Condition | Precision | Recall | F1 |
| --- | --- | --- | --- | --- |
| ncit_doid | 0.9 Threshold | 0.933 | 0.726 | 0.817 |
| omim_ordo | 0.9 Threshold | 0.896 | 0.400 | 0.553 |
| snomed_body_fma | 0.9 Threshold | 0.501 | 0.600 | 0.546 |
| snomed_ncit_neoplasm | 0.9 Threshold | 0.641 | 0.577 | 0.607 |
| snomed_ncit_pharm | 0.9 Threshold | 0.841 | 0.851 | 0.846 |
| **Dataset Average** | **Threshold** | **0.762** | **0.631** | **0.674** |

**Figure 1:**
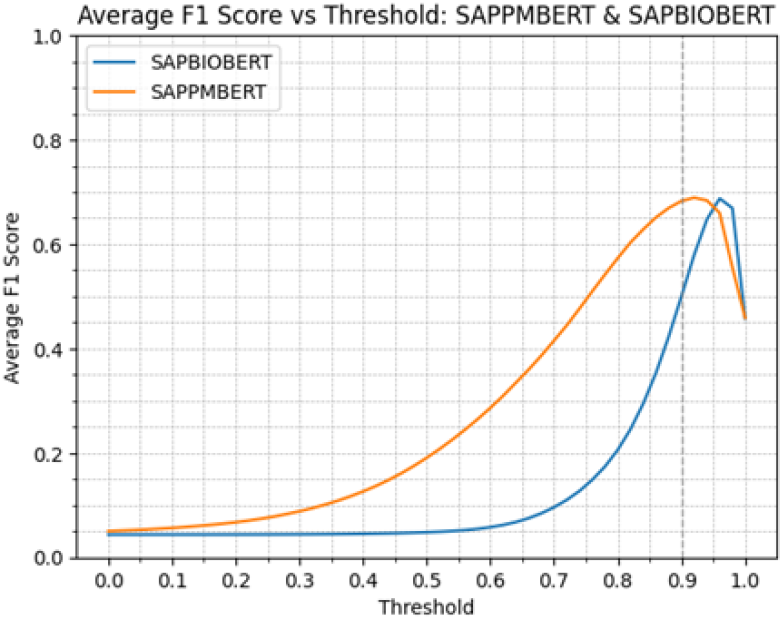
Demonstrating the change in average F1 score across 5 OAEI tasks as the threshold value increases from 0.0 to 1.0 for the two best-performing models

### 4.2 Decoder LLMs Show Mixed Results in Prompting Experiments. Domain-Specific Trained Models Still Outperform Overall

Generative LLMs show mixed results, as can be seen in Table 4. ME-Llama 3 8B underperformed every other model, including a quantised Llama 3.1 8B, hinting at substantive gains between Llama 3.0 and 3.1 at some expense of recall. Both Q4_K_M Llama 3.1 and ME-Llama 3 8B rarely chose “non-match” in multi-choice prompts, showing that this prompt format is too taxing for older or smaller quantised models. Only Q4_K_M Llama 3.3 70B in the true/false no-definition scheme beat the 0.90 heuristic. A Llama 3.1 8B Instruct run failed under the prompting parameters used in this study so its results were not included. This evidence shows that effective prompting techniques vary significantly even within the same model family or company. The domain-finetuned m42-v2 outclassed all Llama 3.1 variants, reinforcing the advantage of medical-specific tuning. LLM-generated differential definitions lifted precision significantly for m42-v2 and Q4_K_M Llama 3.3 70B in both multichoice and true/false prompting. ME-Llama selected a code almost always in the multi-choice task, regardless of the differential definitions. Likewise, Q4_K_M Llama 3.1 ignored added context and seldom returned a non-match. Naturally, in this setting, recall will remain artificially high. Therefore, these models are unreliable for this task. Llama 3.1 8B even appeared to drop in performance with differential definitions, suggesting the task is either too specialised or complex without domain fine-tuning, and comparing this directly to the m42-v2 Llama 3.1 8B suggests that domain fine-tuning is still relevant for performance even for decoder LLMs.

**Table 4:** Evaluation of Decoder Models Across Prompting Conditions. Precision, Recall, and F1 scores are reported for each model across multiple-choice and true/false settings, with and without differential definitions. Results show how prompting strategies affect model performance, with benchmark averages included for reference.

| Model | Condition | Precision | Recall | F1 |
| --- | --- | --- | --- | --- |
| ME-LLaMA 3 8B | MC-Def | 0.427 | 0.614 | 0.501 |
|  | MC-NoDef | 0.394 | 0.762 | 0.519 |
|  | TF-Def | 0.393 | 0.785 | 0.524 |
|  | TF-NoDef | 0.392 | 0.785 | 0.523 |
| m42-V2LLaMA 3.1 8B | MC-Def | 0.575 | 0.666 | 0.616 |
|  | MC-NoDef | 0.452 | 0.763 | 0.568 |
|  | TF-Def | 0.723 | 0.524 | 0.597 |
|  | TF-NoDef | 0.700 | 0.648 | 0.668 |
| LLaMA 3.1 8B | MC-Def | 0.420 | 0.334 | 0.368 |
|  | MC-NoDef | 0.410 | 0.696 | 0.515 |
|  | TF-Def | 0.411 | 0.770 | 0.536 |
|  | TF-NoDef | 0.460 | 0.781 | 0.578 |
| Q4_K_MLLaMA 3.1 8B | MC-Def | 0.364 | 0.727 | 0.485 |
|  | MC-NoDef | 0.370 | 0.740 | 0.493 |
|  | TF-Def | 0.629 | 0.293 | 0.386 |
|  | TF-NoDef | 0.567 | 0.714 | 0.624 |
| Q4_K_MLLaMA 3.3 70B | MC-Def | 0.593 | 0.698 | 0.640 |
|  | MC-NoDef | 0.446 | 0.777 | 0.567 |
|  | TF-Def | 0.726 | 0.619 | 0.665 |
|  | TF-NoDef | 0.650 | 0.719 | 0.679 |
| BenchmarkAverages [3] | OAEI BioML | – | – | 78.62 |
|  | LLMs4OM Best | 61.58 | 56.35 | 58.06 |

## 5 Conclusion

Domain-specific encoder models are still very relevant amongst available open-source AI tools. These models can be lightweight and maintain competitive performance in semantic encoding tasks. This study further demonstrates that domain-specific models outperform their general domain counterparts when adjusting for model size and training data volume. Even when paired with a high-quality RAG, decoder LLMs do not yet meet reliability or performance requirements for fully automated vocabulary standardisation. Progress will likely depend on a few factors. More capable base LLMs may overcome domain-specific gaps, as evidenced by Q4_K_M Llama 3.3 70B outperforming the medical 8B models at the trade-off of 9x the parameters. Massive (Llama 3.3 70B) general-domain releases show encouraging cross-domain gains and may eventually brute force performance. Without extensive contextual scaffolding, multi-choice prompting is still too demanding for today’s readily deployable 8B parameter LLMs and prompting needs to be adjusted to the individual model. A greater availability of high-quality and a greater volume of medical and clinical data may improve the current AI models and assist future development. This study adds to the ongoing challenge that medical domain models face when adopting state-of-the-art AI systems.

## Data Availability

All data and models produced are available online through either open-source or free to use with registration or usage agreement

## Acknowledgments

The authors thank Yashpal Ramakrishnaiah for initial discussions on concept mapping and EHR-QC implementations. This work was supported by computational resources provided by RMIT University. AB acknowledges the STEM PhD scholarship from RMIT University.

## A Sample Prompts

### A.1 Definition Builder Prompt

You are a biomedical ontology expert with deep knowledge of ontologies such as UMLS, SNOMED, and MONDO.

**Context:** You are working with the ontology: `{ontology_name}`. **Term to define:** “`{input_label}`” **Similar labels to distinguish from:** `{Similar Labels}`

**Instructions**

1. Internally reason about how “`{input_label}`” differs from the similar labels.
2. Provide a concise, specific definition for “`{input_label}`” that clearly distinguishes it.
3. Output only the definition text (one paragraph or less), with no additional commentary.
4. Do not include disclaimers or any other text.

**Begin your definition now**:

### A.2 MultiChoice with Differential Definitions

You are an Ontology Matching Expert in UMLS, MONDO, and related ontologies. Below is an input code and several candidate codes with generated definitions from another LLM model. If none matches, respond with None. Otherwise, output only the best label—no explanations.

*Input Details*.

**Code:** `{input_code}`

**Label:** `{input_label}`

**Definition:** `{input_definition}`

*Candidate Matches*. `{items}`

*Instructions*.

1. Compare the input label/definition to each candidate.
2. If one aligns best, output its label exactly; if not, output None.
3. Output *only* that label or None.

*Example: A Suitable Match*.

- Input: Code: 123, Label: “Corneal Disease”, Definition: “A condition affecting the cornea.”
- Candidates: “eye disease”, “corneal dystrophy”
- Output: corneal dystrophy

*Example: No Suitable Match*.

- Input: Code: 321, Label: “Rare Genetic Disorder X”, Definition: “A condition with unique markers.”
- Candidates: “common cold”, “seasonal allergies”
- Output: None

*Answer (Only one label or* None).

### A.3 TrueFalse

You are an Ontology Matching Expert with extensive experience in UMLS, MONDO, and similar ontologies. A previous model generated definitions for each code, highlighting how they may differ within the same ontology. These definitions may be paraphrased or partial.

The two codes below come from different ontologies, but they may represent the same underlying concept. Evaluate their labels and definitions, and respond with yes or no *only* if they truly match.

*Input Concept*.

**Code:** `{input_code}`

**Label:** `{input_label}`

**Definition:** `{input_definition}`

*Candidate Concept*.

**Code:** `{matched_code}`

**Label:** `{matched_label}`

**Definition:** `{matched_definition}`

Do these two concepts refer to the same real-world entity, condition, or idea? Answer yes or no only:

## B Online Resources

- **Machine Learning Friendly OAEI Ontology Dataset** [14]: https://zenodo.org/records/13119437
- **LangChain** [21]: https://www.langchain.com
- **FAISS (Facebook AI Similarity Search)** [27]: https://faiss.ai
- **CHROMA** [8]: https://www.trychroma.com
- **Ollama**[35]: https://ollama.com

